# Collateral damage: the impact on cancer outcomes of the COVID-19 pandemic

**DOI:** 10.1101/2020.04.21.20073833

**Authors:** Amit Sud, Michael Jones, John Broggio, Chey Loveday, Bethany Torr, Alice Garrett, David L. Nicol, Shaman Jhanji, Stephen A. Boyce, Phillip Ward, Jonathan M. Handy, Nadia Yousaf, James Larkin, Yae-Eun Suh, Stephen Scott, Paul D.P. Pharoah, Charles Swanton, Christopher Abbosh, Matthew Williams, Georgios Lyratzopoulos, Richard Houlston, Clare Turnbull

## Abstract

**Background:** Cancer diagnostics and surgery have been disrupted by the response of healthcare services to the COVID-19 pandemic. Progression of cancers during delay will impact on patient long-term survival.

**Methods:** We generated per-day hazard ratios of cancer progression from observational studies and applied these to age-specific, stage-specific cancer survival for England 2013-2017. We modelled per-patient delay of three months and six months and periods of disruption of one year and two years. Using healthcare resource costing, we contextualise attributable lives saved and life years gained from cancer surgery to equivalent volumes of COVID-19 hospitalisations.

**Findings:** Per year, 94,912 resections for major cancers result in 80,406 long-term survivors and 1,717,051 life years gained. Per-patient delay of six months would cause attributable death of 10,555 of these individuals with loss of 205,024 life years. For cancer surgery, average life years gained (LYGs) per patient are 18·1 under standard conditions and 15·9 with a delay of six months (a loss of 2·3 LYG per patient). Taking into account units of healthcare resource (HCRU), surgery results on average per patient in 2·25 resource-adjusted life years gained (RALYGs) under standard conditions and 1·98 RALYGs following delay of six months. For 94,912 hospital COVID-19 admissions, there are 474,505 LYGs requiring of 1,097,937 HCRUs. Hospitalisation of community-acquired COVID-19 patients yields on average per patient 5·0 LYG and 0·43 RALYGs.

**Interpretation:** Delay of six months in surgery for incident cancers would mitigate 43% of life years gained by hospitalisation of an equivalent volume of admissions for community acquired COVID-19. This rises to 62% when considering resource-adjusted life-years gained. To avoid a downstream public health crisis of avoidable cancer deaths, cancer diagnostic and surgical pathways must be maintained at normal throughput, with rapid attention to any backlog already accrued.

**Funding:** Breast Cancer Now, Cancer Research UK, Bobby Moore Fund for Cancer Research, National Institute for Health Research (NIHR)

## INTRODUCTION

Following the first case reports in Hubei province, China in late 2019, a pandemic of COVID-19 coronavirus was declared by the World Health Organisation in March 2020. Whilst COVID-19 causes minimal or mild illness in most, a small but appreciable proportion of individuals require oxygen therapy and often admission to an Intensive Care Unit (ICU). The ensuing unprecedented pressure on hospital wards and ICUs has necessitated rapid repurposing of staff and capacity towards the management of COVID-19 cases with deprioritisation of non-emergency clinical services, including diagnostics and elective specialist surgery. Concurrently, lockdown of the population has led to a reduction in the presentation and referral of symptomatic patients from primary into secondary care.

For many elective procedures, the ensuring delay will impact on quality of life but is unlikely to have long-term consequences. By contrast, for patients with cancer, delay of surgery has the real potential to increase the likelihood of metastatic disease, with some patients’ tumours progressing from being curable (with near normal life expectancy) to non-curable (with limited life expectancy)^1^. The situation has been further exacerbated by recent safety concerns regarding aerosol generation from endoscopy, cystoscopy and surgery^2,3^.

Current projections indicate that COVID-19-related disruption of UK healthcare may well last for 18 months or more, until there is either long term effective containment in the population or large-scale vaccination. To inform healthcare prioritisation and resource allocation during this period requires an evaluation of the consequence of the disruption on other disease-associated mortality. Specifically, we have examined the impact on cancer outcomes of different periods of delay of cancer surgery with disruption extending over variable time periods, comparing resource-weighted outcomes to hospital management of COVID-19 patients. We provide the research, policy community and public with: (i) baseline parameters (surgical case volume, 5-year survival, surgical intervention, resource requirement, pattern of adjuvant chemotherapy) to assist further modelling of cancer services; (ii) an evidence-based model of per-day cancer progression for different tumour types; (iii) initial estimates of attributable deaths and lost actuarial life years for each age-specific, stage-specific group of cancer patients, by per-patient delay and period of disruption; (iv) initial estimates of attributable deaths and lost actuarial life years summed at population-level; (v) using common costings for healthcare staffing, initial comparison of lives saved and actuarial life years gained comparing cancer surgery and hospitalisation of COVID-19 patients.

## METHODS

### Data sources

Number and age-specific 5-year survival of cancer patients that had potentially curative surgical resections for non-haematological malignancies between 2013 and 2017 were obtained from Public Health England National Cancer Registration Service (NCRAS)^4^. As well as cancer stage at diagnosis for each cancer type, breast tumour receptor data allowed subtyping of these cancers as hormone receptor positive (ER+ HER2-), HER+ (any), ER-HER2- and other. Median duration of hospital stay for each cancer type, staffing of theatres, ICU and surgical wards was based on information from three large UK surgical oncology centres. Patterns of administration of adjuvant Systemic Anti-Cancer Therapy (SACT) were based on oncologist-reviewed standard practice guidance^5^. ICU COVID-19 mortality and distribution by age for the UK were obtained from ICNARC^6^. Data from Wuhan was used as the basis of the age distribution of community infection, age-specific likelihoods of admission from community to hospital and mortality rates for non-ICU COVID-19 patients^7,8^. The proportion of hospital patients likely to require ICU was based on data from Lombardy^9^. Requirements for ICU stay and ward day respectively for COVID-19 patients were based on Hospital Admitted Patient Care Activity in 2018-2019 for Viral Pneumonia as well as critical care admission requiring advanced respiratory support^10^.

### Analysis

#### Impact of COVID-associated delay on cancer outcomes

We used published data on the impact on overall survival from delay in cancer surgery to estimate per day hazard ratios (HRs) of mortality associated with delay for the different cancers^11-21^. Where data was not available for a given cancer we assumed equivalence of HRs with a cancer for which an estimated could be generated, based on comparable 5-year survivorship^4^. By accounting for COVID-related post-surgical mortality and changes in SACT, we adjusted five-year survival figures for each cancer for surgical patients under ***standard*** care to estimate ***current*** 5-year survival. To model outcomes of surgery ***post-delay***, we apply to standard 5-year survival, the transition HR for the specified number of days of delay, again including COVID-related post-surgical mortality and changes in SACT. We estimated COVID-associated surgical mortality based on per day rate of nosocomial infection, operation-specific duration of post-surgical admission and age-specific mortality from infection. For cancers routinely treated with SACT, we estimated COVID-19 associated mortality based on per day rate of nosocomial infection, the frequency of SACT scheduling, increased risk associated with immunosuppression and age-specific mortality from infection. Assuming improvement in cold protocols, we modelled different rates of nosocomial between current (10%) and post-delay surgery (5%). In our modelling we assumed that SACT is administered where benefit exceeds COVID-related mortality.

We used actuarial life-expectancies, to calculate life years gained, averaged per patient. We examined reduction in overall survival (OS) and life years gained (LYG), comparing surgery under standard care, current conditions and post-delay, by cancer type and by age and stage. Using 2013-2017 surgical workload data, we calculated across all adult cancers examined, the total number of deaths and life years lost attributable to delay. To address possible scenarios we considered per-patient delay of up to 6-months, and 1- and 2-year periods of disruption.

#### COVID-19 outcome

To compare life years associated with timely cancer surgery with that afforded by hospitalisation of COVID-19 patients, we modelled community-ascertained COVID-19 infections by age, adjusting for demographic differences between England and Wuhan^8,22-24^. We applied age-specific rates of admission of community cases to hospital. We assumed 16% of hospital patients are transferred to ICU^9^. We applied estimates for age-specific survival for ICU patients (ICNARC) and for non-ICU patients using data from Wuhan^7,25^. To calculate attributable life years gained from hospitalisation, we based our analysis on age-specific survival for non-admission to be zero (ICU) and 75% (non-ICU).

#### Resource

We analysed healthcare resource units (HCRU) focused specifically on frontline medical and nursing staff, where one HCRU is one 12-hour shift of direct nursing or medical care. We up-weighted for shifts from healthcare workers of high-salary (senior doctors) and/or of current scarcity (anaesthetists, ICU nurses). We calculated HCRUs per patient using estimated staffing ratios for theatres, ICU and ward care and operation-specific data for theatre hours, ICU stay and ward days from oncology centres.

Details of assumptions and parameter estimates are detailed in **Supplementary Table 1**. Analyses were performed using STATA (version 15) and transcribed to Excel, to provide a full visibility of parametrisation, model outputs and opportunity for the reader to customise parameters (**Supplementary Materials**).

## RESULTS

### Impact of surgical delay on survival for different cancers

The greatest rates of deaths arise following even modest delays to surgery in aggressive cancers, with over 30% reduction in survival at six months and over 17% reduction in survival at three months for patients with stage 2 or 3 cancers of the bladder, lung, oesophagus, ovary, liver, pancreas and stomach (**Table 1, Supplementary Table 2, Supplementary Materials)**. Accounting for nosocomial COVID-19 infection, for cancers with a relatively good overall prognosis, delay of surgery by three months had a minimal impact on survival: <1% for all Stage 1 ER+ and HER2+ breast cancers, for example. In older patients, for tumour types such as prostate cancer and ER+ breast cancer, the current impact on survival of COVID-related mortality exceeded the impact of delay of 3 or even six months (**Table 1, Supplementary Table 2**).

**Table 1:**
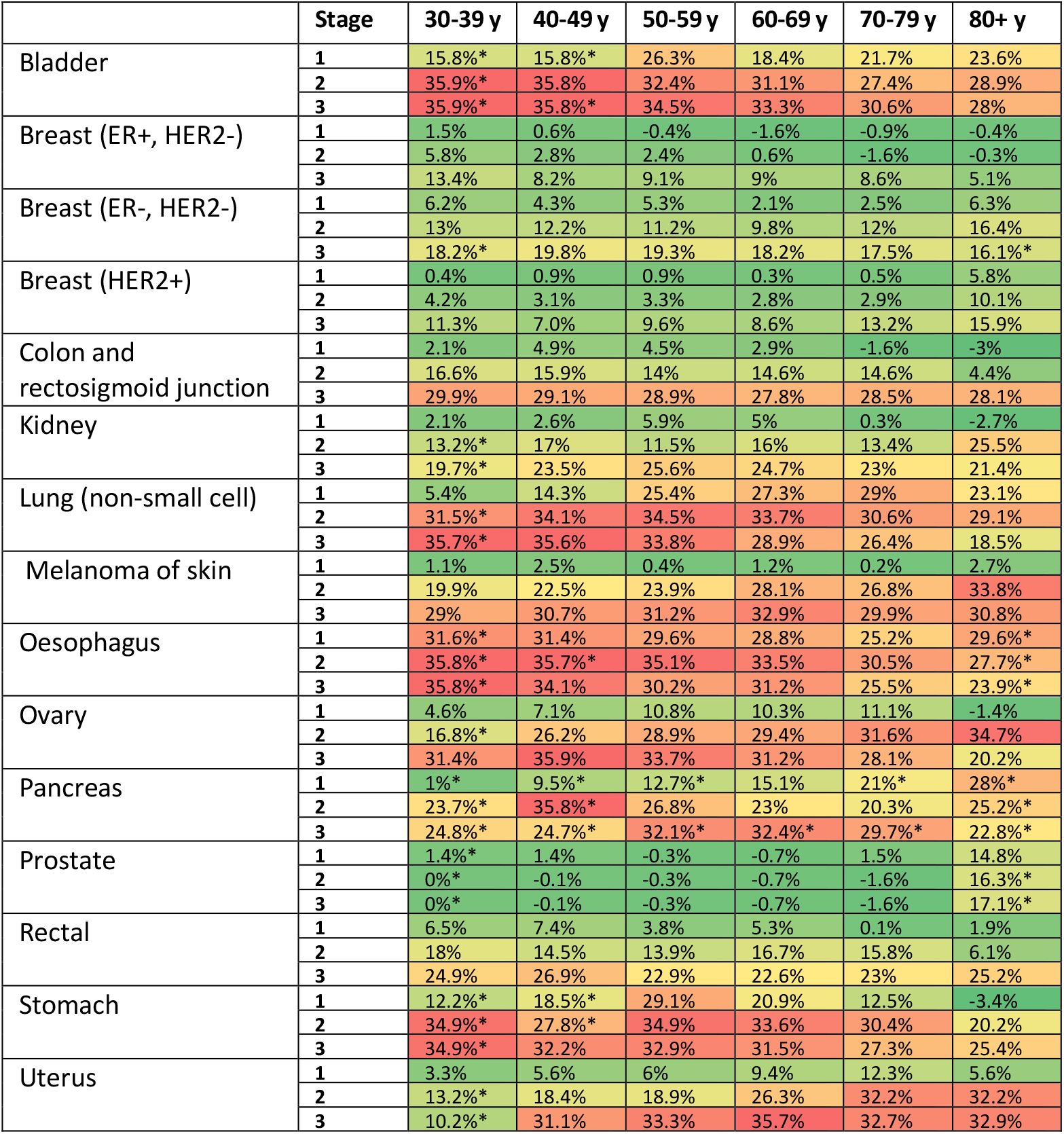
Reduction in survival as a consequence of six-month delay to surgery for 13 cancer types, by tumour stage and age of diagnosis. *indicates strata estimates of lower confidence whereby crude rather than net survival estimates were applied.

|  | Stage | 30-39 y | 40-49 y | 50-59 y | 60-69 y | 70-79 y | 80+ y |
| --- | --- | --- | --- | --- | --- | --- | --- |
| Bladder | 1 | 15.8%* | 15.8%* | 26.3% | 18.4% | 21.7% | 23.6% |
|  | 2 | 35.9%* | 35.8% | 32.4% | 31.1% | 27.4% | 28.9% |
|  | 3 | 35.9%* | 35.8%* | 34.5% | 33.3% | 30.6% | 28% |
| Breast (ER+, HER2-) | 1 | 1.5% | 0.6% | -0.4% | -1.6% | -0.9% | -0.4% |
|  | 2 | 5.8% | 2.8% | 2.4% | 0.6% | -1.6% | -0.3% |
|  | 3 | 13.4% | 8.2% | 9.1% | 9% | 8.6% | 5.1% |
| Breast (ER-, HER2-) | 1 | 6.2% | 4.3% | 5.3% | 2.1% | 2.5% | 6.3% |
|  | 2 | 13% | 12.2% | 11.2% | 9.8% | 12% | 16.4% |
|  | 3 | 18.2%* | 19.8% | 19.3% | 18.2% | 17.5% | 16.1%* |
| Breast (HER2+) | 1 | 0.4% | 0.9% | 0.9% | 0.3% | 0.5% | 5.8% |
|  | 2 | 4.2% | 3.1% | 3.3% | 2.8% | 2.9% | 10.1% |
|  | 3 | 11.3% | 7.0% | 9.6% | 8.6% | 13.2% | 15.9% |
| Colon and rectosigmoid junction | 1 | 2.1% | 4.9% | 4.5% | 2.9% | -1.6% | -3% |
|  | 2 | 16.6% | 15.9% | 14% | 14.6% | 14.6% | 4.4% |
|  | 3 | 29.9% | 29.1% | 28.9% | 27.8% | 28.5% | 28.1% |
| Kidney | 1 | 2.1% | 2.6% | 5.9% | 5% | 0.3% | -2.7% |
|  | 2 | 13.2%* | 17% | 11.5% | 16% | 13.4% | 25.5% |
|  | 3 | 19.7%* | 23.5% | 25.6% | 24.7% | 23% | 21.4% |
| Lung (non-small cell) | 1 | 5.4% | 14.3% | 25.4% | 27.3% | 29% | 23.1% |
|  | 2 | 31.5%* | 34.1% | 34.5% | 33.7% | 30.6% | 29.1% |
|  | 3 | 35.7%* | 35.6% | 33.8% | 28.9% | 26.4% | 18.5% |
| Melanoma of skin | 1 | 1.1% | 2.5% | 0.4% | 1.2% | 0.2% | 2.7% |
|  | 2 | 19.9% | 22.5% | 23.9% | 28.1% | 26.8% | 33.8% |
|  | 3 | 29% | 30.7% | 31.2% | 32.9% | 29.9% | 30.8% |
| Oesophagus | 1 | 31.6%* | 31.4% | 29.6% | 28.8% | 25.2% | 29.6%* |
|  | 2 | 35.8%* | 35.7%* | 35.1% | 33.5% | 30.5% | 27.7%* |
|  | 3 | 35.8%* | 34.1% | 30.2% | 31.2% | 25.5% | 23.9%* |
| Ovary | 1 | 4.6% | 7.1% | 10.8% | 10.3% | 11.1% | -1.4% |
|  | 2 | 16.8%* | 26.2% | 28.9% | 29.4% | 31.6% | 34.7% |
|  | 3 | 31.4% | 35.9% | 33.7% | 31.2% | 28.1% | 20.2% |
| Pancreas | 1 | 1%* | 9.5%* | 12.7%* | 15.1% | 21%* | 28%* |
|  | 2 | 23.7%* | 35.8%* | 26.8% | 23% | 20.3% | 25.2%* |
|  | 3 | 24.8%* | 24.7%* | 32.1%* | 32.4%* | 29.7%* | 22.8%* |
| Prostate | 1 | 1.4%* | 1.4% | -0.3% | -0.7% | 1.5% | 14.8% |
|  | 2 | 0%* | -0.1% | -0.3% | -0.7% | -1.6% | 16.3%* |
|  | 3 | 0%* | -0.1% | -0.3% | -0.7% | -1.6% | 17.1%* |
| Rectal | 1 | 6.5% | 7.4% | 3.8% | 5.3% | 0.1% | 1.9% |
|  | 2 | 18% | 14.5% | 13.9% | 16.7% | 15.8% | 6.1% |
|  | 3 | 24.9% | 26.9% | 22.9% | 22.6% | 23% | 25.2% |
| Stomach | 1 | 12.2%* | 18.5%* | 29.1% | 20.9% | 12.5% | -3.4% |
|  | 2 | 34.9%* | 27.8%* | 34.9% | 33.6% | 30.4% | 20.2% |
|  | 3 | 34.9%* | 32.2% | 32.9% | 31.5% | 27.3% | 25.4% |
| Uterus | 1 | 3.3% | 5.6% | 6% | 9.4% | 12.3% | 5.6% |
|  | 2 | 13.2%* | 18.4% | 18.9% | 26.3% | 32.2% | 32.2% |
|  | 3 | 10.2%* | 31.1% | 33.3% | 35.7% | 32.7% | 32.9% |

For a high proportion of solid cancers, survival at 5 years is generally considered to be equivalent to cure. Predicated on this assertion, we considered gain in actuarial life years gained adjusting for resource, resource adjusted life years (RALYGs). Perhaps not surprisingly, most benefit is afforded in younger age groups for operations that are shorter with no associated ICU requirement. For example, trans urethral resection of stage 1 bladder cancers affords on average 23·4 RALYG per patient age 30-39, whereas cystectomy for stage 2 bladder cancer is only associated with 1·2 RALYGs in that age group (**Supplementary Table 3**). In the context of prioritisation, avoidance of a 6-month delay restitutes on average 4·1 RALYGs in the former group, compared to 0·7 in the latter (**Table 2, Supplementary Table 4**). Wide local excision for breast cancer has low resource requirement and therefore confers substantial RALYGs even in good prognosis subtypes.

**Table 2:** Estimated average life years gained per unit of healthcare resource for cancer surgery, by tumour stage and age of diagnosis for 13 cancer types comparing current surgery to surgery after six months delay. *indicates strata estimates of lower confidence whereby crude rather than net survival estimates were applied.

|  | Stage | 30-39 y | 40-49 y | 50-59 y | 60-69 y | 70-79 y | 80+ y |
| --- | --- | --- | --- | --- | --- | --- | --- |
| Bladder | 1 | 4.11* | 3.28* | 4.14 | 2.02 | 1.48 | 0.83 |
|  | 2 | 0.71* | 0.57 | 0.39 | 0.26 | 0.14 | 0.08 |
|  | 3 | 0.71* | 0.57* | 0.41 | 0.28 | 0.16 | 0.07 |
| Breast (ER+, HER2-) | 1 | 0.31 | 0.1 | -0.04 | -0.14 | -0.05 | -0.01 |
|  | 2 | 1.22 | 0.47 | 0.3 | 0.05 | -0.09 | -0.01 |
|  | 3 | 2.8 | 1.37 | 1.15 | 0.79 | 0.47 | 0.14 |
| Breast (ER-, HER2-) | 1 | 1.29 | 0.72 | 0.67 | 0.19 | 0.14 | 0.18 |
|  | 2 | 2.71 | 2.03 | 1.41 | 0.86 | 0.66 | 0.46 |
|  | 3 | 3.8* | 3.3 | 2.43 | 1.6 | 0.95 | 0.45* |
| Breast (HER2+) | 1 | 0.09 | 0.16 | 0.12 | 0.03 | 0.03 | 0.16 |
|  | 2 | 0.88 | 0.52 | 0.42 | 0.25 | 0.16 | 0.29 |
|  | 3 | 2.36 | 1.16 | 1.2 | 0.76 | 0.72 | 0.45 |
| Colon and rectosigmoid junction | 1 | 0.07 | 0.13 | 0.09 | 0.04 | -0.01 | -0.01 |
|  | 2 | 0.55 | 0.42 | 0.28 | 0.21 | 0.13 | 0.02 |
|  | 3 | 1 | 0.77 | 0.58 | 0.39 | 0.25 | 0.13 |
| Kidney | 1 | 0.14 | 0.13 | 0.23 | 0.14 | 0.01 | -0.02 |
|  | 2 | 0.47* | 0.48 | 0.25 | 0.24 | 0.12 | 0.12 |
|  | 3 | 0.7* | 0.67 | 0.55 | 0.37 | 0.21 | 0.1 |
| Larynx | 1 | 0.36* | 0.41 | 0.36 | 0.22 | 0.09 | 0.08 |
|  | 2 | 0.92* | 0.74* | 0.39* | 0.42 | 0.26 | 0.13* |
|  | 3 | 1* | 0.8* | 0.63 | 0.42 | 0.24 | 0.08* |
| Lung (non-small cell) | 1 | 0.16 | 0.34 | 0.45 | 0.34 | 0.22 | 0.09 |
|  | 2 | 0.93* | 0.8 | 0.62 | 0.42 | 0.24 | 0.12 |
|  | 3 | 1.05* | 0.84 | 0.6 | 0.36 | 0.2 | 0.07 |
| Melanoma of skin | 1 | 0.39 | 0.71 | 0.07 | 0.18 | 0.02 | 0.13 |
|  | 2 | 2.07 | 1.87 | 1.51 | 1.24 | 0.73 | 0.48 |
|  | 3 | 3.03 | 2.56 | 1.96 | 1.45 | 0.81 | 0.43 |
| Oesophagus | 1 | 0.56* | 0.45 | 0.32 | 0.22 | 0.12 | 0.07* |
|  | 2 | 0.64* | 0.51* | 0.38 | 0.25 | 0.14 | 0.07* |
|  | 3 | 0.64* | 0.48 | 0.32 | 0.23 | 0.12 | 0.06* |
| Ovary | 1 | 0.48 | 0.59 | 0.68 | 0.45 | 0.30 | -0.02 |
|  | 2 | 1.76* | 2.18 | 1.82 | 1.3 | 0.86 | 0.49 |
|  | 3 | 0.83 | 0.76 | 0.54 | 0.35 | 0.19 | 0.07 |
| Pancreas | 1 | 0.02* | 0.14* | 0.14* | 0.12 | 0.1* | 0.07* |
|  | 2 | 0.43* | 0.52* | 0.29 | 0.18 | 0.1 | 0.06* |
|  | 3 | 0.45* | 0.36* | 0.35* | 0.25* | 0.14* | 0.06* |
| Prostate | 1 | 0.05* | 0.04 | -0.01 | -0.01 | 0.01 | 0.07 |
|  | 2 | 0* | 0 | -0.01 | -0.01 | -0.02 | 0.08* |
|  | 3 | 0* | 0 | -0.01 | -0.01 | -0.02 | 0.08* |
| Rectal | 1 | 0.22 | 0.2 | 0.08 | 0.07 | 0 | 0.01 |
|  | 2 | 0.6 | 0.39 | 0.28 | 0.23 | 0.14 | 0.03 |
|  | 3 | 0.83 | 0.72 | 0.46 | 0.32 | 0.2 | 0.11 |
| Stomach | 1 | 0.25* | 0.3* | 0.36 | 0.18 | 0.07 | -0.01 |
|  | 2 | 0.67* | 0.42* | 0.4 | 0.27 | 0.15 | 0.05 |
|  | 3 | 0.67* | 0.49 | 0.38 | 0.25 | 0.14 | 0.07 |
| Uterus | 1 | 0.29 | 0.39 | 0.32 | 0.35 | 0.28 | 0.07 |
|  | 2 | 1.15* | 1.28 | 0.99 | 0.97 | 0.73 | 0.38 |
|  | 3 | 0.89* | 2.16 | 1.75 | 1.31 | 0.74 | 0.39 |

### Impact of surgical delay on cancer survival combined across cancer types

Each year, 94,912 surgical resections for common invasive adult cancer types are undertaken in England, with 80,406 of those patients surviving their cancer at 5 years. A surgical delay of three months across all incident solid tumours over one year, would incur 4,471 excess deaths, escalating to 10,555 excess deaths for a six month delay. This includes at six months, attributable deaths of 2,909 for colorectal cancer 1,406 for lung cancer and 804 for breast cancer **(Figure 1)**.

For a high proportion of solid cancers 5-year survival is generally considered to be equivalent to cure. Predicated on this assertion, across all cancers a delay of three months in treatment would lead to a reduction of 87,788 life-years and for six months reduction of 205,024 life years (**Table 3**). **Figure 2** shows the respective long term consequences of delay for different cancer types. Prior to the COVID-19 crisis each year cancer surgery was directly responsible for 1,717,051 LYGs. This represents on average 18·1 LYG per patient, which markedly reduces to 17·2 with three months delay and to 15·9 with six months delay. Cancer surgery per year requires 764,765 units of healthcare resource: assuming this to be unchanged by delay, this affords on average 2·25 RALYG per patient under standard conditions, reducing to 2·13 with three months delay and 1·98 with six months of delay, an average loss of 0.11 and 0.27 RALYGs respectively per patient.

**Table 3:** Summary of England population level outcomes for delay in cancer surgery with comparison to an equivalent number of admissions for community-acquired COVID-19 infection, examining periods of disruption of 12 and 24 months, and per-patient delays of three months and six months.

| Reference period of time/ months |  | 12 | 12 | 24 | 24 |
| --- | --- | --- | --- | --- | --- |
| Cancer Surgery |  |  |  |  |  |
| Per patient delay/ months |  | 3 | 6 | 3 | 6 |
| STANDARD CARE | Major resections for cancer-total | 94,912 |  | 189,823 |  |
|  | HCRUs-total | 764,765 |  | 1,529,529 |  |
|  | LYGs-total | 1,717,051 |  | 3,434,102 |  |
|  | Lives saved-total | 80,406 |  | 160,812 |  |
|  | LYG from cancer treatment- average per patient | 18.09 |  |  |  |
|  | LYG from cancer treatment per HCRU- average per patient | 2.25 |  |  |  |
| IMPACT of DELAY | Deaths attributable to delay-total | 4,471 | 10,555 | 8,943 | 21,111 |
|  | Life years lost attributable to delay-total | 87,788 | 205,024 | 175,577 | 410,049 |
|  | LYG from post-delay cancer treatment- average per patient | 17.17 | 15.93 | 17.17 | 15.93 |
|  | Life years lost attributable to delay- average per patient | 0.92 | 2.16 | 0.92 | 2.16 |
|  | LYG from post-delay cancer treatment per HCRU- average per patient | 2.13 | 1.98 | 2.13 | 1.98 |
|  | Life years lost per HCRU attributable to delay- average per patient | 0.11 | 0.27 | 0.11 | 0.27 |
| Community-acquired COVID-19 management |  |  |  |  |  |
| Community infections |  | 683,083 |  | 1,366,167 |  |
| Hospital Admissions | Total admissions | 94,912 |  | 189,823 |  |
|  | ICU admissions | 15,186 |  | 30,372 |  |
|  | non-ICU admissions | 79,726 |  | 159,451 |  |
| Health care resource units (HCRUs) | Total | 1,097,937 |  | 2,195,875 |  |
|  | ICU | 523,912 |  | 1,047,824 |  |
|  | non-ICU | 574,025 |  | 1,148,051 |  |
| Deaths | Total | 15,141 |  | 30,281 |  |
|  | ICU | 7,756 |  | 15,512 |  |
|  | non-ICU | 7,385 |  | 14,770 |  |
| Lives saved -attributable to hospital admission -total | Total | 25,515 |  | 51,030 |  |
|  | ICU | 7,430 |  | 14,860 |  |
|  | non-ICU | 18,085 |  | 36,170 |  |
| LYGs -attributable to hospital admission -total | Total | 474,505 |  | 949,010 |  |
|  | ICU | 210,096 |  | 420,192 |  |
|  | non-ICU | 264,409 |  | 528,818 |  |
| LYG from hospitalisation- average per patient | All | 5.00 |  |  |  |
|  | ICU | 13.83 |  |  |  |
|  | non-ICU | 3.32 |  |  |  |
| LYG from hospitalisation per HCRU- average per patient | All | 0.43 |  |  |  |
|  | ICU | 0.40 |  |  |  |
|  | non-ICU | 0.46 |  |  |  |

### Resource comparison for outcomes afforded by cancer surgery and COVID-19 management

For contextualisation we compare the impact of cancer surgery delay to hospitalisation for COVID-19 infection. COVID-19 ICU admission for those aged 40-49 yielded on average 27**·**5 LYG and 0**·**8 RALYG. Those aged >80 years admitted to ICU gain on average 2**·**1 LYG and 0**·**06**·**RALYG. For non-ICU admission, average benefit is 9**·**3 LYG and 1**·**3 RALYG for those aged 40-49 and 1**·**4 LYG and 0**·**19 RALYG for those aged >80 years (**Supplementary Materials)**. These estimates are inherently conservative as they do not take into account the impact on life expectancy of the excess comorbidities associated with many hospitalised COVID-19 cases.

COVID-19 community-acquired infection of 683,083 individuals would result in 94,912 hospital admissions (*i*.*e*. the equivalent number to number of annual admissions for cancer surgery). For these 94,912 admissions, 15,186 will require ICU (critical cases) and 79,726 will not require ICU (severe cases). 1,097,937 units of healthcare resource are required in total and there are 15,141 deaths, 25,515 attributable lives saved and 474,505 attributable LYGs (7,756 deaths/7,430 attributable lives saved/210,096 LYGs for ICU admissions, 7,385/ 18,085/ 264,409 for non-ICU). This represents on average 5·00 LYG and 0·43 RALYG per hospitalised COVID-19 patient.

It is therefore noteworthy, that a delay of surgery by six months results in 205,024 lost life years for an annual quota of surgical patients: this equates to 43% of the total 474,505 life years gained from hospitalisation of an equivalent number of community-acquired COVID-19 cases. This rises to 62% when adjusted for differences in resource (RALYGs).

## DISCUSSION

We provide estimates derived from reported surgical outcomes to quantify the impact on survival of delay of cancer treatment. These take into account issues of nosocomial infection specific to the COVID-19 pandemic. We present data for per-patient delays of 3 and six months, and periods of disruption of 1 year and 2 years, but provide a flexible model by which the reader can explore other scenarios (**Supplementary Materials)**. We explore the impact on available life years by modelling 5-year survival to approximate long-term survival and resumption of actuarial life expectancy. To contextualise the impact of delay to cancer surgery against current strains on resource, we compare equivalent admission volumes for cancer surgery and COVID-19 care, examining life years gained against resource required. There are implications of this rapid communication for policymakers, clinicians, and the public.

### Implications for the public

Whilst seeking in general to minimise social contact, prompt presentation to primary care for evaluation and investigation of new symptoms remains essential in order to offer the best opportunity for curative surgery for a cancer. For a significant proportion of patients, delay of 3-six months in diagnosis and surgery will affect the likelihood of achieving long-term survival from their cancer.

### Implications for healthcare planning and resource utilisation

For aggressive cancers, even short delays (three months) have a significant impact on patient survival. However, even for cancers of comparatively favourable prognosis, a moderate delay (six months) will result in significant summed attributable deaths as many of these cancers are common. Delay will also result in tumours being more advanced, meaning not only is survival poorer, but that the upstaged cancers will be more costly to treat both in terms of surgery and/or chemotherapy. Furthermore, resource requirements (for example ICU stay) are dramatically higher for the many who will inevitably present as emergencies such with obstruction, perforation or acute bleeding of the gastrointestinal tract^26^.

Alongside any ‘bulge’ in accumulated cases will be the normal stream of incident cancer presentations. In the face of prolonged stress, it will be challenging to provide extra capacity to address these bulges alongside standard demands. There is a real danger we shall find the backlog from the initial COVID-19 crisis causing knock-on delays that affect delivery of cancer diagnostics and surgery for several years and result in a very substantial excess of deaths.

We need to consider resourcing in the likely event of needing to maintain provision for treating COVID-19 for a sustained period of time, potentially up to 2 years. Although large facilities may be built/repurposed for COVID-19 management, these facilities are competing for the same fixed pool of healthcare workers that provide care for treating non-COVID-19 disease.

Our comparative analyses demonstrate a sizeable drop in cancer survival resulting from short (three month) and moderate (six month) delays in treatment. Delay in cancer surgery will have highly deleterious health and economic impact: for the most part the surgery will still be required (and may be more complex and costly) but results in rapid diminution resultant life years gained and resource-adjusted life years. Comparing equivalent sized hospital populations adjusted for resource, the health impact of delaying cancer surgery for six months will approximate 60% of health gains of hospitalizations for community acquired COVID-19 infection.

### International relevance

While we have used data for England, cancer survival is broadly similar across most economically developed countries, so the impact of delay per tumour is broadly applicable. However, variation in incidence of cancer, actuarial survival and population structure mean that predictions regarding total case numbers and life years gained and lost are more difficult to extrapolate, even when scaling for relative size of reference population.

### Implications for healthcare policy

In the medium-long term (over the next 3-24 months), avoidance of delay to cancer surgery should be of the highest priority: urgent attention is required to ensure sufficient resourcing for standard capacity of cancer diagnostic and surgical pathways. In the short term, to avoid knock-on delays, immediate diversion of supra-normal resource volumes are required to process the backlog of cases that will have accrued in the initial months of the pandemic, in which referrals, investigations and surgeries have been reduced by up to 80%^27^.

Active focus is required to establish ‘cold’ sections of the healthcare system, with rigorous protocols for staff screening and shielding protocols. This will serve to minimise nosocomial acquisition and mortality from COVID-19, to protect staff and also to provide reassurance to the public regarding uptake of diagnostics and surgery for cancer. Urgent review by professional bodies is required regarding best protection of their staffing groups, and guidance on surgical and diagnostic practice commensurate with the true risks^2^.

### Implications for prioritisation amongst cancer patients

Recent guidance issued by NHS England assigned three levels of priority for surgical treatment for non-coronavirus cancer patients: emergency (within 72 hours), urgent (required within 4 weeks to save life/prevent progression of disease beyond operability) and non-urgent (delayed for 10-12 weeks predicted to have no negative outcome)^28^. However, presuming as incorrect the assumption that COVID-related disruption to services will be fully resolved within 12 weeks, nearly all non-emergency cancer cases will by this schema be in the urgent category. Some professional groups have issued more specific tumour-type guidance. Given an accrued backlog of cases and ongoing tight competition for resources, it is inevitable that decisions regarding surgical prioritisation will be required for a number of years, with capacity varying geographically and temporally. Recognising its limitations regarding assumptions and parameters, we propose a model that provides a rational approach by which to evaluate across patients of different ages, tumour types and stages, the benefit and resource implications of their cancer surgery. We highlight in our model those age-stage groups for which COVID-related mortality currently exceeds survival benefit for surgery and/or SACT. Whilst these and other groups for whom benefit is marginal will be the most rationale to delay, they will nevertheless require monitoring and surgery downstream. Longitudinal planning, monitoring of progression, dynamic re-prioritisation and capacity-planning will inevitably be highly challenging.

### Limitations

As with any model-based analysis, our predictions are predicated on the validity of assumptions and estimates used for parameterisation. While we have made use of observational data, our approach simplifies the complexity of cancer progression and is solely survival-focused. For healthcare planning, a more elaborate model capturing stage-shifting may offer additional utility. The benefit of SACT is simplistic as the dosing, benefits and immunosuppressive consequences vary by chemotherapy regimen. Mortality from nosocomial COVID-19 infection during surgical admission or attendance for chemotherapy is based on a uniform per-day risk of infection: these may vary between institutions. Our model of COVID-19 admissions is limited by availability of detailed individual-level UK data, in particular for non-CCU hospital admissions, thus necessitating reliance on data from China and Italy. While our resourcing analysis deliberately focuses on the requirement for the direct medical and nursing staff who most limit healthcare provision, we acknowledge it does not capture other ‘costs’ incurred in hospital care, primary care and social care.

### Further research

Within our current approach we only estimate the effects of a specified period of per-patient delay. Data from NHS activity over the last 2 months offer the prospect of developing dynamic models to predict the impact of (i) differential prioritisation of patient groups, (ii) different patterns of re-presentation of ‘accumulated’ cases alongside incident cases and (iii) varying release of bottlenecks in primary care, diagnostics and surgery. Evaluation is also important for the alternative management approaches being adopted, such as radiotherapy with curative intent where surgery is gold-standard or *a priori* hormonal treatment for prostate and ER-positive breast cancers. For any strategies involving deliberate delay to surgery, models for re-staging and dynamic re-prioritisation are essential. We have focused on the impact to surgery with curative intent; analyses are also required to quantify the impact on mortality of withholding of life-extending chemo- and radio-therapy for patients with Stage 4 disease.

We have only compared resource-adjusted outcomes between cancer surgery and hospitalisation of community-acquired COVID-19 infection. Systematic resource-adjusted outcome analysis across all aspects of healthcare would serve to inform competing priorities. More broadly, assuming there will be universal contraction in available resources for all non-COVID-19 healthcare, health-economic re-evaluation of cancer care protocols will bee required as availability of treatments of high cost per QALY are likely to require review.

## CONCLUSION

Decisions regarding prioritisation and ring-fencing of healthcare capacity need to be evidence-based. To ensure that the health benefits of hospitalisation for community-acquired COVID-19 infection are not ultimately dwarfed by the consequences of delayed management for patients with cancer and other life-threating diseases, rapid comparative impact analyses such as ours are required.

Compared to COVID-19 management, cancer surgery is highly impactful in regard of life-years gained per resource expended. Delay in diagnosis and surgery cause exponential burden of attributable mortality. The COVID-19 pandemic has placed unprecedented strain on health care provision; it is highly plausible that this pressure will continue for up to two years. Supra-normal capacity is required when addressing backlogs of accumulated cancer cases alongside ongoing incident cases. To avoid a deferred public health crisis of unnecessary cancer deaths, urgent ringfencing of substantial resources is required.

## Data Availability

This work uses data that has been provided by patients and collected by the NHS as part of their care and support. The data are collated, maintained and quality assured by the National Cancer Registration and Analysis Service, which is part of Public Health England (PHE).

## LEGENDS FOR TABLES AND FIGURES

**Figure 1:** Attributable deaths in England over one year from per-patient delay to surgery of six months, for all solid cancers analysed and six common cancer types.

**Figure 2:** Life years lost in England over one year from per-patient delay to surgery of six months, for all solid cancers analysed and six common cancer types.

## Author contributions

C.T., M.E.J., A.S. and R.S.H. designed the model. M.E.J. provided cancer progression models. J.B. generated and quality-assured the NCRAS datasets applied to the model. M.E.J., J.B. C.T., R.S.H., A.S., C.A., G.L., M.W., and P.D.P.P provided epidemiological expertise in parameterisation of the model. S.A.B, S.J., D.L.N, P.W., J.L., J.M.H, N.Y. and Y-E.S provided details of clinical pathways and estimates of clinical resourcing. B.T., A.G. and C.L. quality assured and user-tested the model. C.L and B.T. assembled figures for presentation. C.T drafted the manuscript, with substantial contribution from A.S., R.S.H., M.E.J., G.L., M.W. and C.S.. All authors contributed to the final manuscript.

## Acknowledgements

MEJ additionally received funding from Breast Cancer Now. GL is supported by a Cancer Research UK Advanced Clinician Scientist Fellowship Award [C18081/A18180] and is Associate Director of the multi-institutional CanTest Collaborative funded by Cancer Research UK [C8640/A23385]. B.T and A.G. are supported by Cancer Research UK award C61296/A27223. R.S.H. is supported by Cancer Research UK (C1298/A8362) and Bobby Moore Fund for Cancer Research UK). A.S. is in receipt of an Academic Clinical Lectureship from National Institute for Health Research (NIHR) and Biomedical Research Centre (BRC) post-doctoral support. This is a summary of independent research supported by the NIHR BRC at the Royal Marsden NHS Foundation Trust and Institute of Cancer Research. The views expressed are those of the authors and not necessarily those of the NHS, NIHR or the Department of Health. This work uses data that has been provided by patients and collected by the NHS as part of their care and support. The data are collated, maintained and quality assured by the National Cancer Registration and Analysis Service, which is part of Public Health England (PHE).

## RESEARCH IN CONTEXT

### Evidence before this study

Coronavirus disease 2019 (COVID-19) has resulted in disruption of elective medical services, including diagnostics and surgery for cancer. When delivered, outcomes from cancer treatment will be influenced by nosocomial COVID-19 infection. Various observational studies, with inherent confounding by indication, have studied for different tumour types the impact of delay to diagnosis on long-term survival. There has been no systematic evaluation of the impact on survival of global delay to surgery for individual tumour types or summed across tumour types. There has been no standardised analysis of survival comparing hospitalisation for community acquired COVID-19 and cancer surgery.

### Added value of this study

To our knowledge, we provided the first explicit analysis of the impact on long-term survival of different per-patient delays to surgery. Combining detailed data on cancer surgery in England 2013-2017 from the National Cancer Registration Analysis Service with per-day hazard ratios of cancer progression from observational studies, we quantified the impact of different periods of per-patient delay on cancer survival, both for age-specific stage-specific individual tumour groups, and summed at population level for different durations of disruption. We found delay of just three months caused substantial impact on survival for many tumour groups. Adjusted for resource, the reduction in total survival resultant from a sixth-month per patient delay to cancer surgery approaches the magnitude of all survival gain from hospitalisation of an equivalent number of community-acquired COVID-19 infections.

### Implications of all the available evidence

A substantial back-log of cancer cases has already accrued, on account of reluctance of patients to seek healthcare, disruption to primary care, diversion of staff from cancer diagnostics and surgery and concern about the safety of these procedures. Supranormal capacity will be required to address swiftly the accumulated back-log. Failure to ring-fence capacity both for this back-log and for maintaining of rapid pathways for cancer diagnosis and surgery, will result in a sizeable delayed public health crisis of avoidable cancer deaths.

